# Drug-Target Mendelian Randomization and Imaging Mediation Analyses Reveal Therapeutic Targets and Causal Mechanisms for Cardiomyopathies

**DOI:** 10.64898/2026.04.20.26351344

**Authors:** Pusu Wang, Yang Song, Bin Zhang, Jixin Yang

## Abstract

**Background:** Hypertrophic (HCM) and dilated (DCM) cardiomyopathy constitute the principal phenotypes of primary cardiomyopathy, yet both lack sufficient therapeutic options. Integrating genetic insights with detailed cardiac phenotyping offers a promising strategy to prioritize targets and elucidate their mechanisms of action.

**Methods:** We conducted an three-stage analysis. First, drug-target Mendelian randomization (MR) was performed using *cis*-acting protein (pQTL) and expression (eQTL) quantitative trait loci as genetic instruments for potential drug targets. Second, we examined causal associations between 82 cardiac magnetic resonance (CMR)-derived imaging traits and HCM/DCM risk in a CMR-based MR analysis. Third, mediation MR was employed to quantify the proportion of the genetic effect of prioritized drug targets on cardiomyopathy risk that was mediated through specific CMR phenotypes.

**Results:** Our analyses identified 19 and 13 potential therapeutic targets for HCM and DCM, respectively. CMR-based MR revealed that HCM risk was causally associated with increased right ventricular ejection fraction (RVEF) and greater left ventricular wall thickness, whereas DCM risk was linked to ventricular dilation, impaired myocardial strain, and altered aortic dimensions. Critically, mediation analysis established that these CMR traits served as significant intermediate pathways. The protective effect of *ALPK3* on HCM risk was mediated through a reduction in myocardial wall thickness. Conversely, the effects of *PDLIM5, HSPA4*, and *FBXO32* on DCM risk were exerted in part via alterations in aortic dimensions.

**Conclusion:** This integrative genetic and imaging study systematically identify candidate therapeutic targets for HCM and DCM and delineates the specific CMR phenotypes through which they likely exert their causal effects. Our findings advance the understanding of disease pathogenesis and highlight new possibilities for improving the diagnosis and management of cardiomyopathy.

## Background

Cardiomyopathy represents one of the most severe outcomes in pediatric and young adult’s heart disease and serves as the primary reason for heart transplantation in children older than one year of age^1,2^. Among these, hypertrophic cardiomyopathy (HCM) and dilated cardiomyopathy (DCM) are two major phenotypic extremes that impose significant clinical and economic challenges. HCM, affecting approximately 1 in 200–500 individuals globally, is characterized by disproportionate myocardial hypertrophy without chamber dilation, often presenting with left ventricular outflow tract obstruction and arrhythmia risk^3,4^. In contrast, DCM, with a prevalence of 1 in 250, manifests as progressive left ventricular dilation and systolic dysfunction, frequently leading to heart failure and sudden cardiac death^5^. These conditions exhibit stark differences in clinical presentation: HCM patients typically present with exertional dyspnea, chest pain, or syncope due to dynamic obstruction, while DCM patients often exhibit fatigue, edema, and signs of systemic congestion from impaired contractility^6^.

Pathologically, HCM stems predominantly from mutations in genes encoding sarcomere proteins (such as MYBPC3 and MYH7), leading to asymmetric hypertrophy^7^. In contrast, DCM has a more heterogeneous genetic basis involving variants affecting sarcomeric, nuclear envelope, and cytoskeletal proteins, ultimately driving ventricular dilation and contractile failure^6^. These distinct pathogenic mechanisms are reflected in differing diagnostic and therapeutic paradigms. Current diagnosis relies heavily on imaging, particularly echocardiography and cardiac magnetic resonance (CMR). For HCM, recent studies underscore the importance of regular CMR-based risk assessment and the adoption of targeted therapies like mavacamten, a cardiac myosin inhibitor that mitigates outflow obstruction^8,9^. For DCM, CMR offers superior characterization of myocardial structure, tissue composition (e.g., fibrosis, edema, inflammation, or storage diseases), and thus plays a critical role in risk stratification for arrhythmias and heart failure^5^. Management of DCM, however, remains anchored in guideline-directed medical therapy for heart failure (e.g., ACE inhibitors, beta-blockers), with few interventions specifically designed to modify the underlying disease^10^. Despite these advances, important gaps persist. The comparative effectiveness of pathophysiology-specific treatment strategies for HCM versus DCM has not been clearly established. While targeted agents like myosin inhibitors are emerging for HCM, DCM care still relies largely on nonspecific heart failure regimens, and true disease-modifying therapies are scarce. Furthermore, evidence-based protocols for pediatric cardiomyopathy are lacking, and CMR remains underutilized in routine practice^11,12^.

In this context, integrating genetic insights offers a promising avenue to refine diagnostic precision and therapeutic targeting. Mendelian randomization (MR) analysis, which leverages genetic variants as instrumental variables, provides a robust framework for inferring causal relationships between drug targets and disease outcomes. Coupled with CMR traits, MR can help elucidate the role of specific molecular pathways in cardiomyopathy pathophysiology and assess their potential as therapeutic targets. Thus, combining drug-target MR with detailed CMR-based phenotyping may bridge critical gaps in our understanding of HCM and DCM, potentially advancing more precise diagnostic and treatment strategies for cardiomyopathies.

## Methods

### Study Design

The overall study design, outlined in Figure 1, consisted of three analytical stages. First, in the drug-targeted MR analysis, we utilized cis-pQTLs and cis-eQTLs to systematically investigate circulating proteins and gene expressions as potential therapeutic targets for hypertrophic and dilated cardiomyopathy. Second, the CMR-based MR analysis was employed to examine potential causal relationships between 82 CMR imaging-derived traits and the risks of HCM and DCM. Third, to elucidate underlying mechanistic pathways, we performed mediation MR analyses to assess whether the identified effects of candidate drug targets on cardiomyopathy were mediated through specific CMR trait.

**Figure 1.**
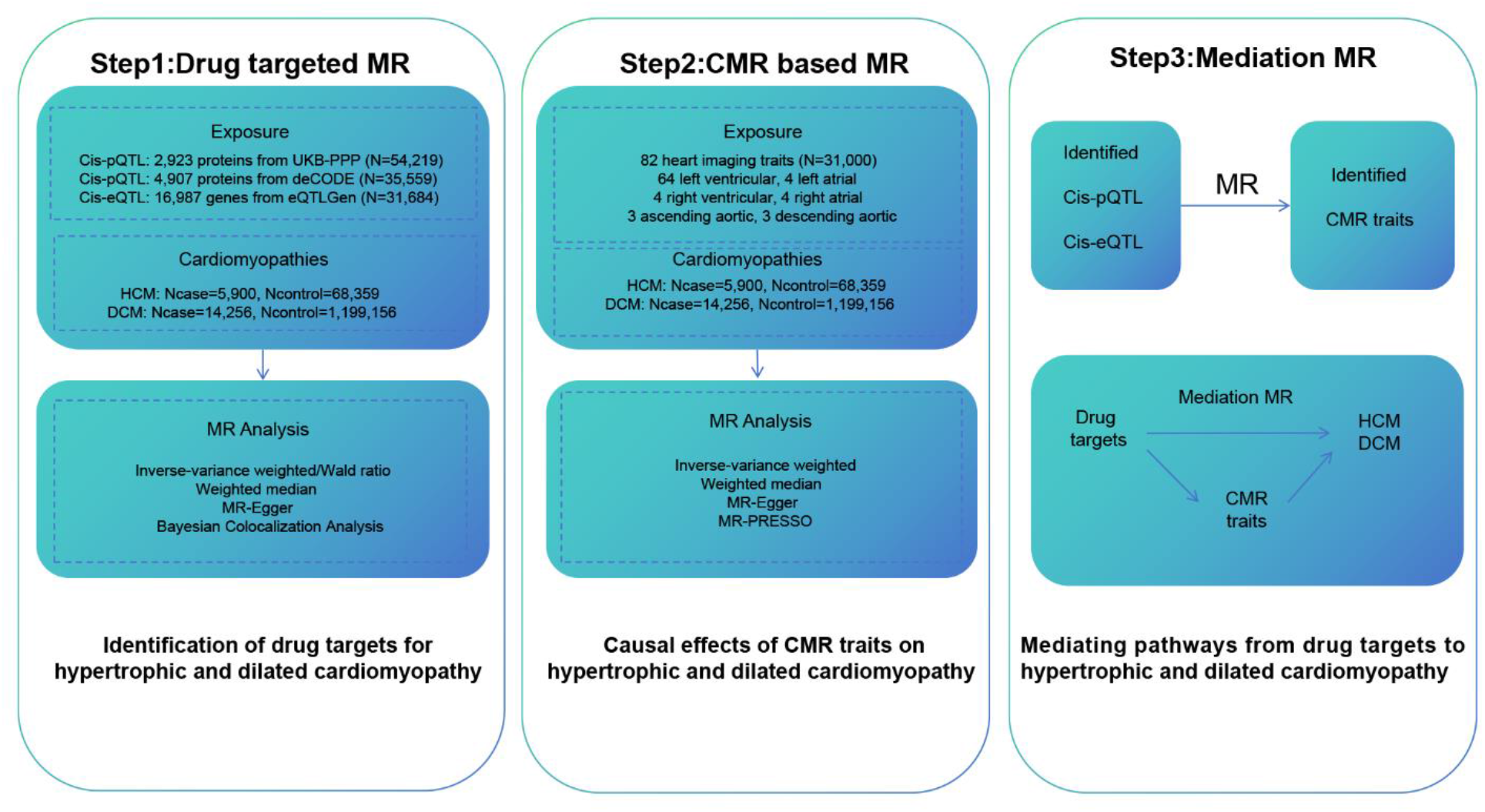
An overview of the study design. CMR, cardiac magnetic resonance; Cis-eqtl, cis-expression quantitative trait loci; Cis-pqtl, cis-protein quantitative trait loci; DCM, dilated cardiomyopathy; HCM, hypertrophic cardiomyopathy; MR, Mendelian randomization.

### Data Sources for Instrumental Variables

Genetic instruments for plasma protein levels were derived from two large-scale proteomic studies: the deCODE study (N=35,559; 4,907 proteins)^13^ and the UK Biobank Pharma Proteomics Project (UKB-PPP, N=54,219; 2,923 proteins)^14^. Selection of valid cis-pQTL instruments adhered to five sequential criteria: (i) genome-wide significant association with protein abundance (P < 5 × 10^−8^); (ii) independence ensured via linkage disequilibrium (LD) clumping (window: 10,000 kb; r^2^ < 0.001); (iii) physical location within ±1000 kb of the encoding gene’s transcription start site; (iv) instrument strength requirement (F-statistic > 10) to minimize weak instrument bias; and (v) exclusion of SNPs associated with the outcome (P < 5 × 10^−5^) to limit potential violation of MR assumptions. Blood-derived expression quantitative trait loci (eQTL) data were obtained from the eQTLGen consortium (31,684 samples; cis-eQTLs for 16,987 genes)^15^. Included cis-eQTLs were required to show a genome-wide significant association with gene expression (P < 5 × 10^−8^), possess a minor allele frequency > 0.01, demonstrate sufficient strength (F-statistic > 10), and map within 1000 kb of the corresponding gene.

Genetic instruments for CMR traits were sourced from a GWAS of 82 heart imaging phenotypes in the UK Biobank British cohort (N=31,000)^16^. These phenotypes comprised 64 left ventricular, 4 left atrial, 4 right ventricular, 4 right atrial, 3 ascending aortic, and 3 descending aortic traits. To ensure adequate SNP coverage for MR analysis, the significance threshold for instrument selection was set at P < 5 × 10^−6^ for these traits. Subsequent filtering for independence, strength, and outcome association mirrored the procedures described for pQTLs.

### Data Sources for Outcomes

To minimize bias from population stratification, primary analyses utilized the largest available GWAS summary statistics for DCM and HCM from predominantly European ancestry populations. HCM data came from a meta-analysis of seven studies (5,900 cases; 68,359 controls; ~92.5% European descent)^17^. DCM data were obtained from the HERMES Consortium (14,256 cases; 1,199,156 controls; primarily European ancestry)^18^. All data used in this study are publicly accessible. The contributing original studies had received appropriate ethics approvals from their respective institutional review boards. Further details regarding dataset characteristics and sources are summarized in Supplementary Table S1 and S2.

### MR and Mediation Analysis

For primary causal inference, we employed the inverse-variance weighted (IVW) method as the main analytical approach, supplemented by the Wald ratio method^19^. Specifically, the Wald ratio was used for exposures instrumented by a single genetic variant, while the IVW method was applied when two or more independent instrumental variables were available. Associations with a false discovery rate (FDR)-adjusted p-value < 0.05 were considered statistically significant evidence supporting a potential causal relationship. To assess the robustness of the primary IVW estimates, we performed complementary analyses using the weighted median and MR-Egger regression methods. The weighted median method provides consistent estimates under the condition that at least 50% of the information comes from valid instruments^20^, while MR-Egger regression can detect and adjust for directional pleiotropy^21^. To provide more robust causal estimates, the MR-PRESSO method was additionally applied as a complement to the primary IVW analysis in our CMR-based MR^22^. We conducted several sensitivity analyses to evaluate the validity of our causal inferences. Heterogeneity among instrumental variable estimates was assessed using Cochran’s Q statistic^23^. In the presence of significant heterogeneity (Q-test p-value < 0.05), the inverse-variance weighted method with multiplicative random effects was employed to provide more conservative and robust causal estimates by allowing for variability in the causal effects across genetic variants^24^. The presence of directional pleiotropy was formally tested by examining the intercept term in the MR-Egger regression, where a statistically significant intercept indicates unbalanced horizontal pleiotropic effects across genetic instruments^22^.

A mediation MR framework was applied to examine whether the effect of a prioritized drug target on cardiomyopathy risk was mediated through specific CMR imaging traits. First, we estimated the causal effect of a candidate drug target (identified from drug targeted MR analysis) on a candidate CMR trait. To qualify for formal mediation testing, three prerequisite associations all had to demonstrate statistical significance after FDR correction (adjusted P < 0.05): (1) the drug target with cardiomyopathy, (2) the CMR trait with cardiomyopathy, and (3) the drug target with the CMR trait. For qualifying trios, the mediation analysis was conducted. The mediated (indirect) effect was calculated as the product of the two path coefficients: β(A), representing the causal effect of the drug target on the CMR trait, and β(B), representing the causal effect of the CMR trait on the cardiomyopathy (i.e., Indirect Effect = β(A) × β(B)). The total causal effect of the drug target on cardiomyopathy was obtained from the initial univariable MR analysis. The proportion of the total effect mediated by the CMR trait was then derived using the formula: (Indirect Effect / Total Effect) × 100%. To account for the uncertainty in both path estimates, the 95% confidence interval for this mediation proportion was calculated using the delta method. This analytical approach allows for the quantification of how much a specific imaging-derived physiological pathway contributes to the overall causal relationship between a drug target and cardiomyopathy^25^. R software (version 4.5.2) was employed to perform all statistical analyses.

### Sensitivity Analyses

Causal inferences were evaluated for both robustness and statistical significance. Robustness required consistent effect directions from the primary IVW estimator and sensitivity analyses (weighted median, MR-Egger, MR-PRESSO), alongside no significant signal of horizontal pleiotropy (assessed via the MR-Egger intercept test). Statistical significance was defined by a FDR-adjusted P-value of less than 0.05. Associations meeting both sets of criteria were considered reliable.

### Bayesian Colocalization Analysis

To evaluate whether the genetic associations for pQTLs and eQTLs with HCM and dilated DCM were driven by shared causal variants, we performed Bayesian colocalization analyses. Specifically, we conducted pairwise colocalization tests between: (1) pQTL and HCM/DCM GWAS summary statistics^26^, and (2) eQTL and HCM/DCM GWAS summary statistics within the corresponding genomic regions^27^. This approach calculates the posterior probability for five distinct hypotheses regarding the genetic architecture underlying two traits in a given locus. Our primary focus was on Hypothesis 4 (PPH4), which posits that both the molecular trait (protein or gene expression) and the cardiomyopathy phenotype are associated with the region due to a single shared causal variant^28^. A posterior probability for Hypothesis 4 (PPH4 > 0.8) was considered evidence of potential colocalization, suggesting a common genetic origin for the observed associations.

## Results

### Potential Drug Targets and Bayesian Colocalization Analysis

The results of drug targeted MR are shown in Figure 2, Fig. S1 - S4, Table S3 - S10. Following MR and sensitivity analyses, we identified significant associations for 8 cis-pQTLs with HCM and 3 with DCM (FDR-P < 0.05). Notably, the proteins CST1 (cystatin-SN, OR = 0.81; 95% CI: 0.73–0.90) and SEMA3G (semaphorin-3G, OR = 0.65; 95% CI: 0.52–0.81) from deCODE, alongside CALCA (calcitonin, OR = 0.35; 95% CI: 0.23–0.54), EPHB6 (ephrin type-B receptor 6, OR = 0.76; 95% CI: 0.66– 0.88), and LATS1 (serine/threonine-protein kinase LATS1, OR = 0.20; 95% CI: 0.10– 0.39) from UKB-PPP, were associated with reduced HCM risk. Conversely, GIPC2 (PDZ domain-containing protein GIPC2, OR = 1.45; 95% CI: 1.23–1.71), LRRC37A2 (leucine-rich repeat-containing protein 37A2, OR = 1.24; 95% CI: 1.14– 1.34), and SPON1 (spondin-1, OR = 1.40; 95% CI: 1.21–1.63) from UKB-PPP were associated with increased HCM risk. For DCM, DNAJB4 (DnaJ homolog subfamily B member 4, OR = 0.72; 95% CI: 0.61–0.84) from deCODE and GIPC2 (OR = 0.81; 95% CI: 0.74–0.90) from UKB-PPP were protective, while LATS1 from UKB-PPP conferred risk (OR = 2.80; 95% CI: 1.79–4.38). Colocalization analysis further supported a shared causal variant for HCM risk with the pQTLs of SEMA3G, CALCA, GIPC2, and SPON1, and for DCM risk with DNAJB4 and GIPC2 (all PP.H4.abf > 0.8), nominating these proteins as potential therapeutic targets.

**Figure 2.**
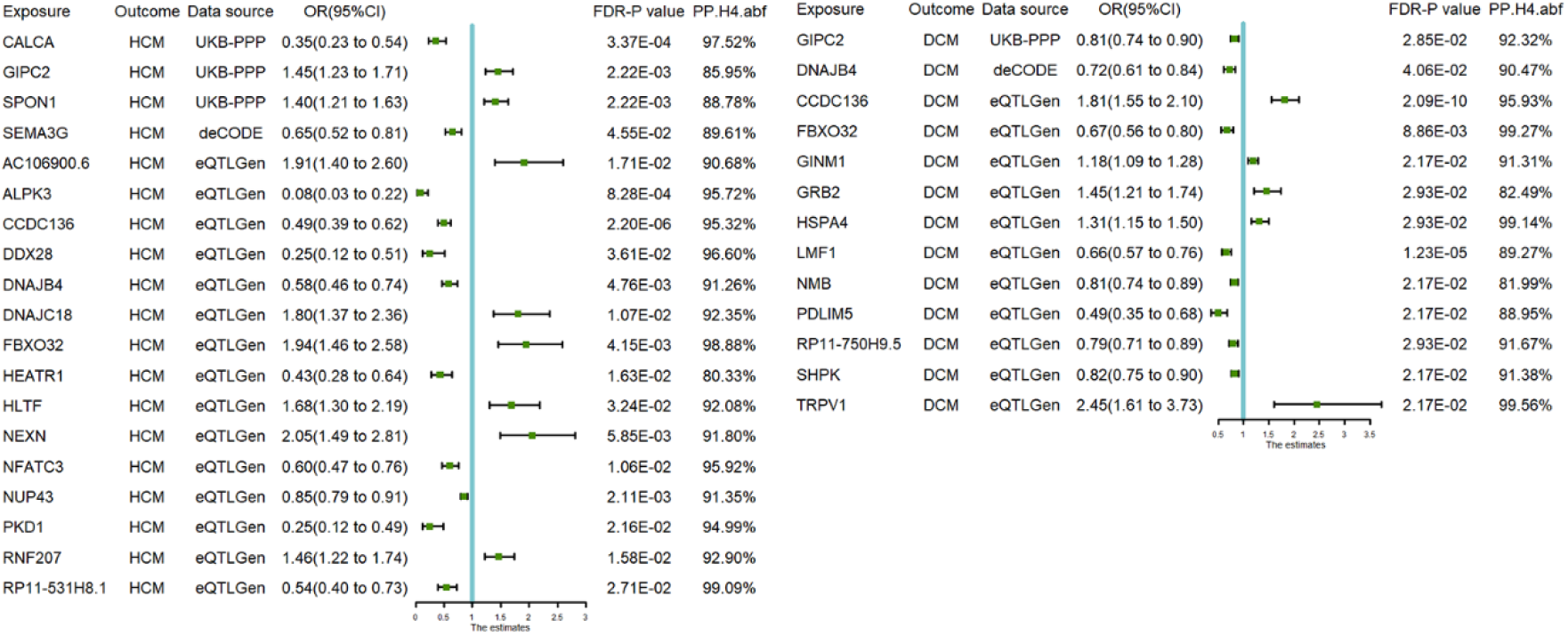
Causal estimates of potential drug targets for HCM and DCM. Cis-eQTLs and cis-pQTLs with an FDR-P value < 0.05 and a PPH4 > 0.8 were considered potential drug targets. ALPK3, alpha-protein kinase 3; CCDC136, coiled-coil domain-containing protein 136; DCM, dilated cardiomyopathy; DNAJB4, DnaJ homolog subfamily B member 4; DNAJC18, DnaJ homolog subfamily C member 18; DDX28, probable ATP-dependent RNA helicase DDX28; FDR, false discovery rate; FBXO32, F-box only protein 32; GINM1, glycoprotein integral membrane protein 1; GRB2, growth factor receptor-bound protein 2; HCM, hypertrophic cardiomyopathy; HEATR1, HEAT repeat-containing protein 1; HLTF, DNA-dependent ATPase/E3 ubiquitin-protein ligase HLTF; HSPA4, heat shock 70 kDa protein 4; LMF1, lipase maturation factor 1; NEXN, nexilin; NFATC3, nuclear factor of activated T-cells, cytoplasmic 3; NMB, neuromedin-B; NUP43, nucleoporin Nup43; OR, odds ratio; PDLIM5, PDZ and LIM domain protein 5; PKD1, polycystin-1; PP.H4.abf, posterior probability of hypothesis 4 (based on approximate Bayes factor); RNF207, RING finger protein 207; SHPK, sedoheptulokinase; TRPV1, transient receptor potential cation channel subfamily V member 1.

Summary-data-based Mendelian randomization analysis revealed 53 cis-eQTLs significantly associated with HCM and 30 with DCM. Colocalization analysis provided high-confidence support (PP.H4.abf > 0.8) for a shared causal mechanism between the disease risk loci and the eQTLs of 15 genes for HCM and 11 genes for DCM. The spectrum of effect sizes for the identified cis-eQTLs varied substantially between the two cardiomyopathy subtypes. For HCM, the odds ratios (ORs) spanned from 0.08 (95% CI: 0.03–0.22) for Alpha-protein kinase 3 (*ALPK3*), indicating a strong protective association, to 2.05 (95% CI: 1.49–2.81) for Nexilin (*NEXN*), a known risk-increasing gene. Similarly, the effects for DCM ranged from an OR of 0.49 (95% CI: 0.35–0.68) for the protective gene PDZ and LIM domain protein 5 (*PDLIM5*) to 2.45 (95% CI: 1.61–3.73) for Transient receptor potential cation channel subfamily V member 1 (*TRPV1*).

### CMR based MR

The CMR-based analysis identified four CMR traits significantly associated with HCM and eleven traits significantly associated with DCM (Figure 3 and Table S11 - S13). For HCM, increased disease risk was significantly linked with right ventricular ejection fraction (RVEF), greater global myocardial-wall thickness at end-diastole (WT global), and greater wall thickness in two specific regional segments at end-diastole (WT AHA 5 and WT AHA 6). The corresponding ORs ranged from 1.78 (95% CI 1.31, 2.42) for RVEF to 2.05 (95% CI 1.38, 3.06) for WT global. For DCM, increased risk was significantly associated with larger left ventricular end-diastolic volume (LVEDV), larger right ventricular end-systolic volume (RVESV), greater ascending aorta maximum area (Aao max area), smaller descending aorta minimum area (Dao min area), impaired global peak circumferential strain (Ecc global), and impaired regional peak circumferential strain in five segments (Ecc AHA 2, 7, 8, 10, and 15). In contrast, higher left ventricular ejection fraction (LVEF) was significantly associated with reduced DCM risk. The ORs for these traits spanned from 0.61 (95% CI 0.47, 0.80) for LVEF to 1.70 (95% CI 1.33, 2.16) for Ecc AHA 8.

**Figure 3.**
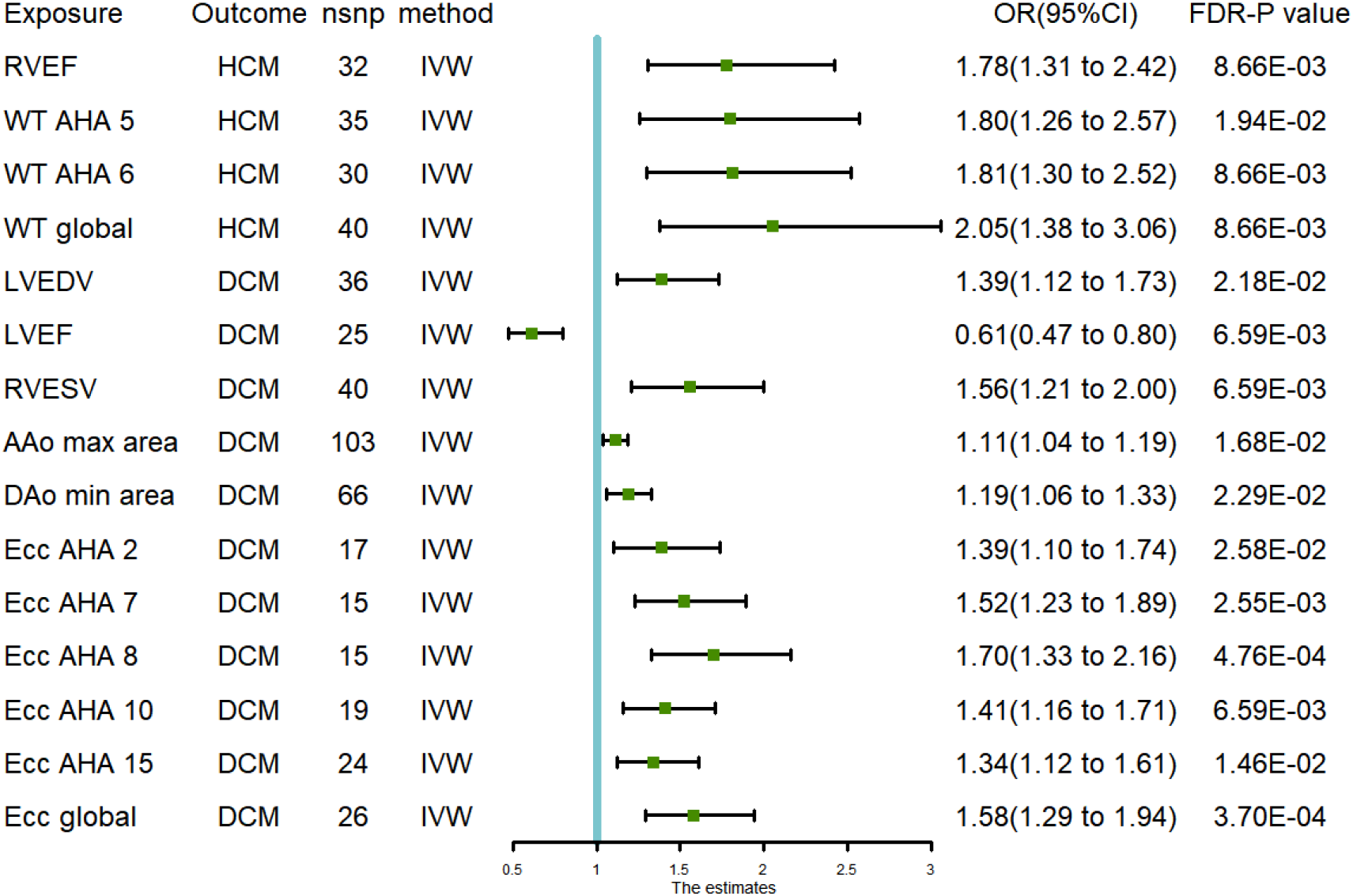
Significant Association of CMR Traits with HCM and DCM. Aao max area, ascending aorta maximum area; Dao min area, descending aorta minimum area; DCM, dilated cardiomyopathy; Ecc AHA, regional peak circumferential strain; Ecc global, global peak circumferential strain; FDR, false discovery rate; HCM, hypertrophic cardiomyopathy; IVW, inverse-variance weighted; LVEDV, left ventricular end-diastolic volume; LVEF, left ventricular ejection fraction; OR, odds ratio; RVEF, right ventricular ejection fraction; RVESV, right ventricular end-systolic volume; WT AHA, regional myocardial-wall thickness at end-diastole; WT global, global myocardial-wall thickness at end-diastole.

### Mediation analysis

MR analysis was performed using cardiomyopathy-associated cis-pQTLs and cis-eQTLs as instrumental variables for cardiomyopathy-related CMR traits, revealing 4 and 28 significant causal associations for cis-pQTLs and cis-eQTLs, respectively (Table S14 - 17). Subsequent mediation analysis delineated six distinct pathways through which cardiomyopathy-related CMR traits mediated the effects of cis-eQTLs on cardiomyopathy susceptibility (Figure 4 and Table S18). Notably, no significant mediating pathways were identified for cis-pQTLs. Specifically, *ALPK3* was found to reduce the risk of HCM by decreasing wall thickness in specific segments (WT AHA 5: 17.03%; WT AHA 6: 18.46%) and globally (WT global: 22.60%). For DCM, *PDLIM5* exerted a protective effect by reducing the descending Dao min area (6.75%), whereas *HSPA4* (encoding Heat shock 70 kDa protein 4) increased DCM risk by augmenting the same trait (8.42%). Additionally, *FBXO32* (F-box only protein 32) appeared to confer protection by decreasing the ascending Aao max area (4.80%). Collectively, these findings indicate that alterations in cardiac structure and function may represent critical intermediary mechanisms for genetic effects on cardiomyopathy, providing a rationale for targeting these pathways in future drug development and management strategies.

**Figure 4.**
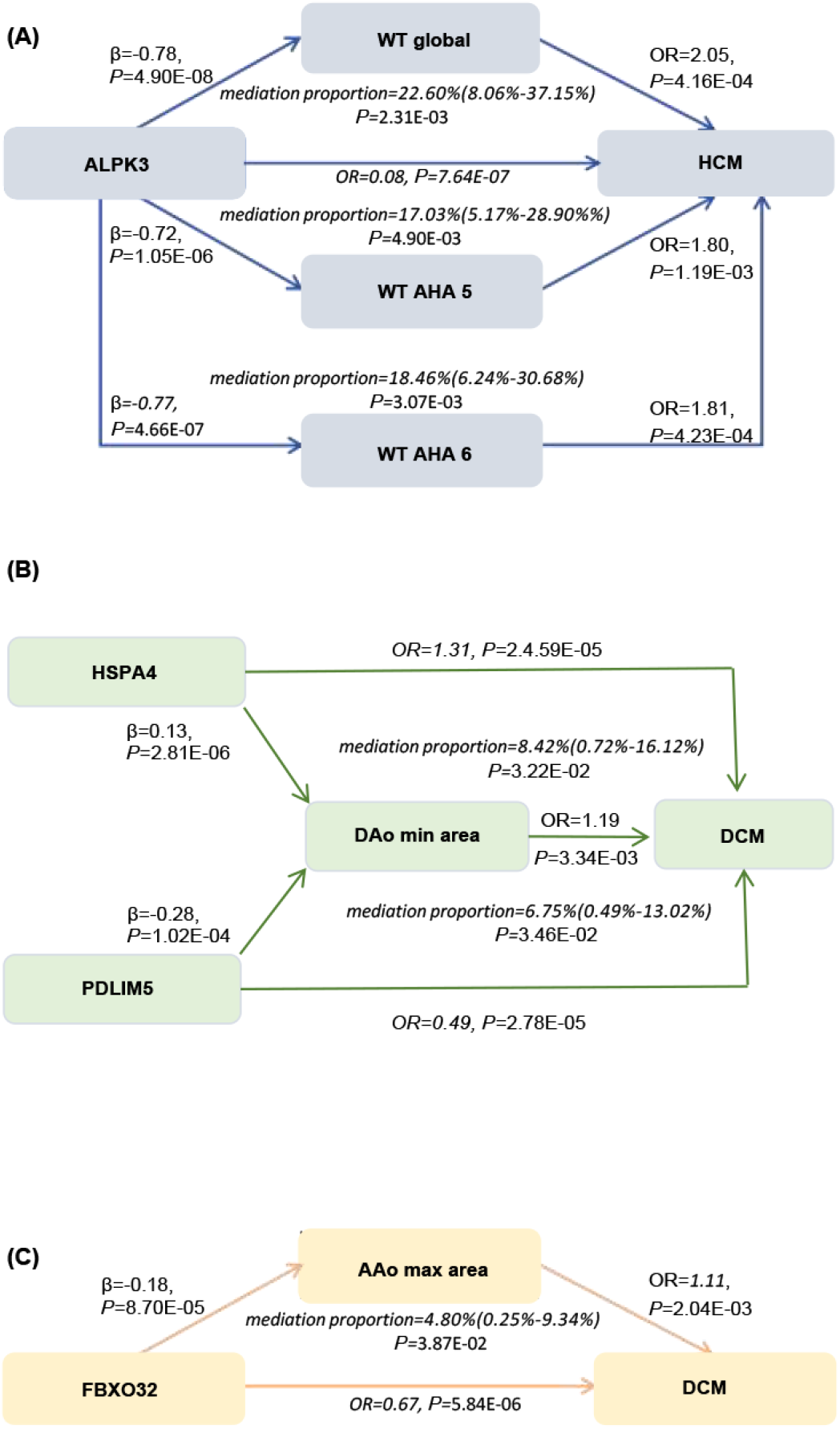
Mediating roles of CMR traits in the causal association between identified drug targets and outcomes (HCM, DCM). Aao max area, ascending aorta maximum area; ALPK3, alpha-protein kinase 3; Dao min area, descending aorta minimum area; DCM, dilated cardiomyopathy; FBXO32, F-box only protein 32; HCM, hypertrophic cardiomyopathy; HSPA4, heat shock 70 kDa protein 4; OR, odds ratio; PDLIM5, PDZ and LIM domain protein 5; WT AHA, regional myocardial-wall thickness at end-diastole; WT global, global myocardial-wall thickness at end-diastole.

## Discussion

This study systematically investigated the causal pathways through which genetic instruments (cis-pQTLs and cis-eQTLs) influence cardiomyopathy risk via cardiac imaging phenotypes by integrating drug-target MR, CMR phenotyping, and a mediation MR framework. Furthermore, we have delineated specific pathways through which key genes mediate their effects via quantifiable cardiac imaging phenotypes. These discoveries deepen our understanding of the pathophysiology of cardiomyopathies, particularly HCM and DCM, and provide novel perspectives for targeted therapy and precision management.

The MR identified a more extensive array of candidate drug targets via cis-eQTLs (15 for HCM, 11 for DCM) compared to cis-pQTLs (4 for HCM, 2 for DCM). Importantly, several of these prioritized proteins and genes are supported by existing clinical or basic research in cardiomyopathy, lending biological credibility to our findings and highlighting promising therapeutic avenues. Our cis-pQTL MR analysis identified SPON1 as being significantly associated with an increased risk of HCM. This aligns with a prospective proteomic study which proposed SPON1 as a potential biomarker for the development of atrial fibrillation in HCM patients, suggesting its involvement in the disease’s pathophysiology^29^. From the cis-eQTL analysis, *ALPK3* and *PKD1* were found to be significantly associated with a reduced risk of HCM. This is supported by mechanistic evidence: heterozygous truncating variants in *ALPK3* are a known cause of autosomal dominant HCM, characterized by a distinct phenotype of concentric or apical hypertrophy and extensive fibrosis^30^. Furthermore, activation of the *PKD1* signaling pathway has been shown to prevent maladaptive cardiac hypertrophy in models of lipid overload^31^. Regarding DCM, our *cis*-eQTL analysis indicated a protective role for *FBXO32*, which aligns with reports that homozygous *FBXO32* mutations drive severe ventricular dilation, systolic dysfunction, and the accumulation of autophagy-related proteins in patients^32^. The convergence of our MR findings with these established mechanistic studies significantly strengthens the reliability of candidate therapeutic targets. However, a portion of the targets identified in our analysis currently lack robust prior biological validation. This underscores an important gap in knowledge and highlights the need for dedicated functional studies to confirm their roles in cardiomyopathy pathogenesis and assess their therapeutic potential.

CMR has become indispensable for the comprehensive phenotyping and risk assessment of cardiomyopathies, providing unparalleled insights into biventricular structure, function, and tissue composition. Our analysis identifies specific CMR-derived metrics significantly associated with an elevated risk for HCM, including an increased RVEF, greater global left ventricular myocardial wall thickness at end-diastole (WT global), and localized thickening of the WT AHA 5 and WT AHA 6 segments. In asymptomatic carriers of pathogenic sarcomeric variants, elevated RVEF may represent an early, pre-clinical CMR biomarker^33^. The pathophysiological cascade in HCM, initiated by mutations that induce a hypercontractile state and activate profibrotic pathways like TGFβ, alongside microvascular dysfunction, ultimately manifests as the characteristic hypertrophy, myocyte disarray, and replacement fibrosis precisely quantified by CMR^7,34^. Conversely, for DCM, increased risk is associated with larger LVEDV and RVESV, altered aortic dimensions, and impaired global and regional peak circumferential strain, while a higher LVEF is protective. CMR serves as the diagnostic gold standard for DCM, accurately quantifying systolic function, ventricular remodeling, and fibrosis. The underlying pathophysiology, whether genetic or acquired, converges on sarcomeric dysfunction and energetic deficit, leading to the eccentric remodeling and systolic failure captured by CMR^35^. Thus, beyond definitive diagnosis, CMR-based tissue and strain characterization provides critical prognostic stratification, guides therapeutic decisions, and offers potential surrogate endpoints for evaluating novel therapies across the spectrum of cardiomyopathies.

This study identifies a protective role for *ALPK3*, wherein it reduces HCM risk by decreasing myocardial wall thickness, both globally (22.60% mediation for WT global) and in specific basal segments (17.03% for WT AHA 5; 18.46% for WT AHA 6). This finding aligns with cohort studies establishing heterozygous truncating variants in *ALPK3* (ALPK3tv) as a significant cause of autosomal dominant HCM, accounting for 1.5–2.5% of cases. ALPK3tv carriers present a distinct phenotype of apical/concentric hypertrophy, extensive late gadolinium enhancement on CMR, and shortened PR intervals. Critically, they harbor a significantly elevated heart failure risk compared to sarcomere-negative HCM patients, with 9% experiencing events during a median 5.3-year follow-up^30^. The underlying pathomechanism involves ALPK3, a muscle-specific pseudokinase localized to the nuclear envelope and sarcomeric M-band. Its loss disrupts cardiac integrity via dual pathways: impaired force buffering due to mislocalization of key proteins (e.g., MYOM1/MYOM2) and dysregulated sarcomere protein turnover via altered M-band-associated proteolysis (e.g., reduced MuRF1, elevated CAPN3). This explains the bimodal phenotypic spectrum—recessive mutations causing infantile dilated cardiomyopathy via structural failure, while heterozygous variants drive compensatory hypertrophy in adulthood^36^. Therapeutic exploration remains nascent. The myosin inhibitor mavacamten, used in obstructive HCM, showed mixed effects in ALPK3tv (K201X) cardiomyocytes, partially correcting diastolic calcium and function but reducing fractional shortening. However, its clinical efficacy remains unclear^37^. Conversely, gene therapy using MyoAAV-miniALPK3 in Alpk3-/-mice successfully restored ALPK3 M-band localization, normalized cardiac structure and function, and markedly improved long-term survival. These preclinical studies confirm *ALPK3* cardiomyopathy is targetable^38^. Our mediation analysis identified left ventricular wall thickness as a key modifiable endpoint for targeted therapy in ALPK3-associated HCM.

Our mediation analysis further elucidated distinct genetic contributions to DCM risk through aortic remodeling traits. Specifically, *PDLIM5* exerted a protective effect by reducing the minimum area of the descending aorta, while *HSPA4* increased DCM risk by augmenting this same trait. In addition, *FBXO32* appeared to confer protection by decreasing the maximum area of the ascending aorta. These findings align with established molecular mechanisms. *PDLIM5 (ENH)* deficiency disrupts Z-line integrity by destabilizing the ENH-CypherS-Calsarcin-1 complex, which leads to Z-line widening and contractile impairment. This results in the progressive left ventricular dilation and systolic dysfunction that hallmark DCM^39^. *HSPA4* expression is upregulated in both pressure-overloaded murine hearts and failing human hearts. Genetic ablation of Hspa4 in mice induced cardiac hypertrophy and fibrosis while preserving contractile function, a phenotype that delineates its specific role in regulating myocardial structural remodeling^40^. Homozygous mutations in *FBXO32* trigger early-onset dilated cardiomyopathy by inducing ER-stress-mediated apoptosis. This pathogenic cascade involves upregulation of CHOP and impaired ubiquitination of ATF2, which collectively disrupt cardiac proteostasis, promote cardiomyocyte loss, and drive ventricular remodeling^41^. Currently, DCM management remains largely reliant on guideline-directed systemic heart failure therapies, with few interventions directly targeting the underlying disease mechanisms. Our analysis, which maps specific genetic variants to quantifiable aortic and structural phenotypes, provides a theoretical foundation for developing more targeted therapeutic strategies in DCM.

Several limitations of this study should be acknowledged. First, our analyses relied predominantly on summary-level data from large-scale biobanks and consortia comprising individuals of primarily European ancestry. This limits the generalizability of our findings to other ancestral populations, where differences in genetic architecture, linkage disequilibrium patterns, and disease prevalence may influence the associations. Second, since both the genetic instruments for certain drug targets and the CMR traits were derived from the UK Biobank, partial sample overlap may introduce bias and inflate statistical associations. Finally, although this study identifies priority candidate targets and plausible mechanistic pathways, it does not provide functional validation. The therapeutic potential of targets such as FBXO32 or HSPA4, along with the clinical utility of the proposed CMR traits, must be prospectively evaluated in experimental models and clinical trials.

## Conclusion

This study integrates drug-target MR with detailed CMR phenotyping to systematically explore therapeutic targets and elucidate causal pathways in HCM and DCM. Our analyses identified 19 potential drug targets along with 4 associated CMR traits for HCM, and 13 drug targets with 11 associated CMR traits for DCM. Furthermore, we found that genes such as *ALPK3, PDLIM5, HSPA4*, and *FBXO32* exert their effects on cardiomyopathy risk through specific CMR traits. These findings may advance our understanding of disease pathogenesis and offer new possibilities for the diagnosis and management of cardiomyopathy, though they await validation in future studies.

## Acknowledgments

Sincere appreciation is expressed to every participant and all investigators involved in the incorporated GWAS projects. We are also indebted to the original study authors for generously making the necessary GWAS summary-level data publicly available.

## Abbreviations

Aao: Ascending aorta
ALPK3: Alpha-protein kinase 3
CMR: Cardiac magnetic resonance
DCM: Dilated cardiomyopathy
Dao: Descending aorta
DNAJB4: DnaJ homolog subfamily B member 4
Ecc: Peak circumferential strain
eQTL: Expression quantitative trait locus
FBXO32: F-box only protein 32
FDR: False discovery rate
GWAS: Genome-wide association study
HCM: Hypertrophic cardiomyopathy
HSPA4: Heat shock 70 kDa protein 4
IVW: Inverse-variance weighted
LD: Linkage disequilibrium
LVEDV: Left ventricular end-diastolic volume
LVEF: Left ventricular ejection fraction
MR: Mendelian randomization
OR: Odds ratio
PDLIM5: PDZ and LIM domain protein 5
pQTL: Protein quantitative trait locus
RVEF: Right ventricular ejection fraction
RVESV: Right ventricular end-systolic volume
SNP: Single nucleotide polymorphism
UKB-PPP: UK Biobank Pharma Proteomics Project
WT: Wall thickness

## Author Contributions

PS. W, JX. Y conceived, designed and drafted the manuscript. PS. W analyzed the data and prepared Figures and tables. JX. Y, B.Z and Y. S made contributions to drafted and revised the manuscript. All authors contributed to the article and approved the submitted version.

## Data Availability Statement

All GWAS summary statistics used in this study can be found in the cited references. Custom analysis code will be shared upon a reasonable request to the corresponding author.

